# Exploring subthreshold functional network alterations in women with phenylketonuria by higher criticism

**DOI:** 10.1101/2024.09.16.24313700

**Authors:** Benedikt Sundermann, Reinhold Feldmann, Christian Mathys, Stefan Garde, Johanna M. H. Rau, Anke McLeod, Josef Weglage, Bettina Pfleiderer

**Author notes:** Correspondence to: Prof. Dr. rer. nat. Dr. med. Bettina Pfleiderer, Clinic of Radiology, Medical Faculty, University of Münster, Albert-Schweitzer-Campus 1, Building A1, 48149 Münster, Germany.

## Abstract

**Objective:** Phenylketonuria (PKU) is an inherited disorder of amino acid metabolism. Despite early dietary treatment, cognitive functioning of patients has been reported as being inferior to healthy controls. Objective of this study was to assess functional connectivity (FC) alterations in PKU in cognition-related brain networks by resting-state functional magnetic resonance imaging. We followed a hierarchical analysis approach partially based on higher criticism (HC) statistics as previously applied in a larger sister-project in fetal alcohol syndrome.

**Results:** After exclusions for excessive head movement, 11 female young adults with early-treated PKU (age: 27.2 ± 3.7 years) and 11 age-matched female healthy controls (age: 25.9 ± 3.8 years) were included in the analysis. We observed effects within attention networks and the default mode network, but not in fronto-parietal networks, at the HC-based intermediate analysis level. No between-network FC differences were found. In the most detailed analysis level, we could not identify single affected functional connections. Despite statistical power limitations in this small sample, findings are in line with previously reported FC alterations in PKU and the cognitive profile in young adults with PKU, particularly with the still uncertain notion that cognitive control deficits might become less pronounced when PKU patients reach adulthood.

## Introduction

Classical phenylketonuria (PKU, OMIM 261600, https://www.omim.org/entry/261600) is an inherited metabolic disease affecting neurocognitive development [1-3]. If treated early by dietary intervention, most affected individuals reach normal or near-normal cognitive abilities [1, 2]. However, many PKU patients exhibit minor cognitive deficits, involving executive functions [4-7]. A recent meta-analysis of cognitive outcomes in adults with early treated PKU observed the most pronounced impairments in reasoning, visual-spatial attention, sustained attention, visuo-motor control, and flexibility [8].

Neuroimaging studies in early-treated PKU have mainly focused on structural alterations [9]. These include diffuse T2w white matter hyperintensities (WMH), predominantly in parieto-occipital regions, but also locally reduced cortical thickness and alterations to white matter tracts [9]. An anterior-posterior gradient has been observed in such morphometric studies [10, 11], suggesting a potential association with the spatial distribution (posterior dominance) of WMH. Functional neuroimaging studies in PKU, including functional magnetic resonance imaging (fMRI) have focused on task-based protocols examining higher cognitive functions [12-16]. Knowledge about cerebral alterations from a functional network perspective is, however, still very limited. Christ et al. observed decreased functional connectivity in the default mode network (DMN) by resting state fMRI (rs-fMRI) in a small and very heterogeneous PKU sample [17]. Potential DMN alterations could also be indirectly inferred from interpretations of altered activity patterns in task-based fMRI [12]. Further evidence of altered functional connectivity (FC) or network dysfunction in PKU results from a study in task-based FC of the prefrontal cortices [14]. Another rs-fMRI study is ongoing [18].

Reliably assessing and interpreting functional neuroimaging data in diseases like PKU is challenging. Generally, sample sizes in neuroimaging studies in affected individuals are limited by the rarity of this inherited metabolic disorder affecting neurocognitive development [1, 9], similar to other neurodevelopmental disorders [19]. This scarcity of data points limits the detectability of disease-related alterations in conventional fMRI analyses, which commonly employ mass-univariate statistical testing [20]. Statistical power limitations in such studies also impair the reliability of published findings [21-24]. In the aforementioned research scenario, few strongly localized statistically significant findings might frequently not well represent more distributed underlying effects at the level of functional networks [20, 21]. Therefore, complementary methods taking wider patterns of activation or connectivity estimates (both above and below conventional statistical thresholds) into account might be better suited to elucidate robust network alterations in the aforementioned patient groups. Among several approaches to explore subthreshold effects in fMRI, analyses using higher criticism (HC) statistics have been suggested [20]. HC is a method to identify rare and weak effects in high-dimensional data [25-27]. It follows the rationale of p-value histogram interpretation to detect an excess of low p-values in a range of primary tests in order to reject a global null hypothesis [28].

We recently reported the application of multi-scale FC modelling based on HC statistics in fetal alcohol syndrome (FAS): This approach identified subnetworks containing FC group differences while single altered connections could not be reliably identified using conventional analysis methods [29]. Here, we applied this analysis approach to a substantially smaller sample of young women with PKU. The small sample size, resulting from necessary exclusions for excessive head motion in the MRI scanner [30] and the rarity of PKU [1], would not be regarded as sufficient for conventional rs-fMRI analyses. We therefore consider this to be exploratory work.

The main goal of this study was to examine FC in cognition-related functional brain networks in young adults with early-treated PKU. The following hypotheses were tested, hereby conceptually following the approach previously reported [29]: (1) FC in the connectome of all brain regions constituting cognition-related brain networks is altered in young adults with PKU compared with a healthy control group. (2) FC within individual cognition-related brain networks is altered in PKU compared with healthy controls (network-wise global hypotheses and individual connections). (3) FC between cognition-related brain networks is altered in PKU compared with healthy controls (global hypothesis and individual connections).

## Materials and Methods

This study was part of a larger functional neuroimaging project in PKU and FAS with overlapping samples [12, 29, 31]. PKU-related aspects of this study follow a concomitant task-based fMRI study [12], and fMRI methodology follows the related study in FAS [29] within this project. Related project details are thus only briefly summarized here.

### Subjects

Primary inclusion and exclusion were identical with the task-based fMRI study in PKU and have been published there [12]. The exclusion criterion of participation in an earlier task-based fMRI study on inhibitory control in males with PKU [13] eventually limited the sample to women, since no sufficiently large group of male subjects with PKU could be recruited. 17 patients attended the MRI examination. Complete MRI data could be acquired in 15 patients. After data acquisition, further subjects were excluded after review of potentially biasing psychoactive medication, structural brain lesions, and MRI data quality control before HC-based group analyses (criteria identical with the accompanying rs-fMRI study in FAS [29]): Data from further 4 PKU patients were excluded because of excessive head motion (2 based on quantitative criteria, 2 based on visual quality control criteria / carpet plots, see below). All results are based on the remaining 11 PKU subjects. All PKU patients received early dietary treatment according to the then-current recommendations of the German working group on pediatric metabolic disorders [32].

An identical number (n = 11) of female healthy controls were drawn from the control group in the PKU/FAS neuroimaging research project [12, 29, 31] by age-based matching. This was necessary since the control group was on average older than the PKU group. Control subjects fulfilled the same inclusion and exclusion criteria as the PKU group, except the primary diagnosis.

Questionnaires included the Edinburgh Handedness Inventory (EHI) [33] and a trail-making task (TMT) of processing speed [34]. Intelligence quotients (IQ) were estimated based on these TMT results [34, 35]. Statistical tests on clinical and demographical data were carried out in SPSS (version 27.0, IBM, Armonk, NY, USA, RRID:SCR_002865).

Demographical data and test results are presented in Table 1.

**Table 1.**
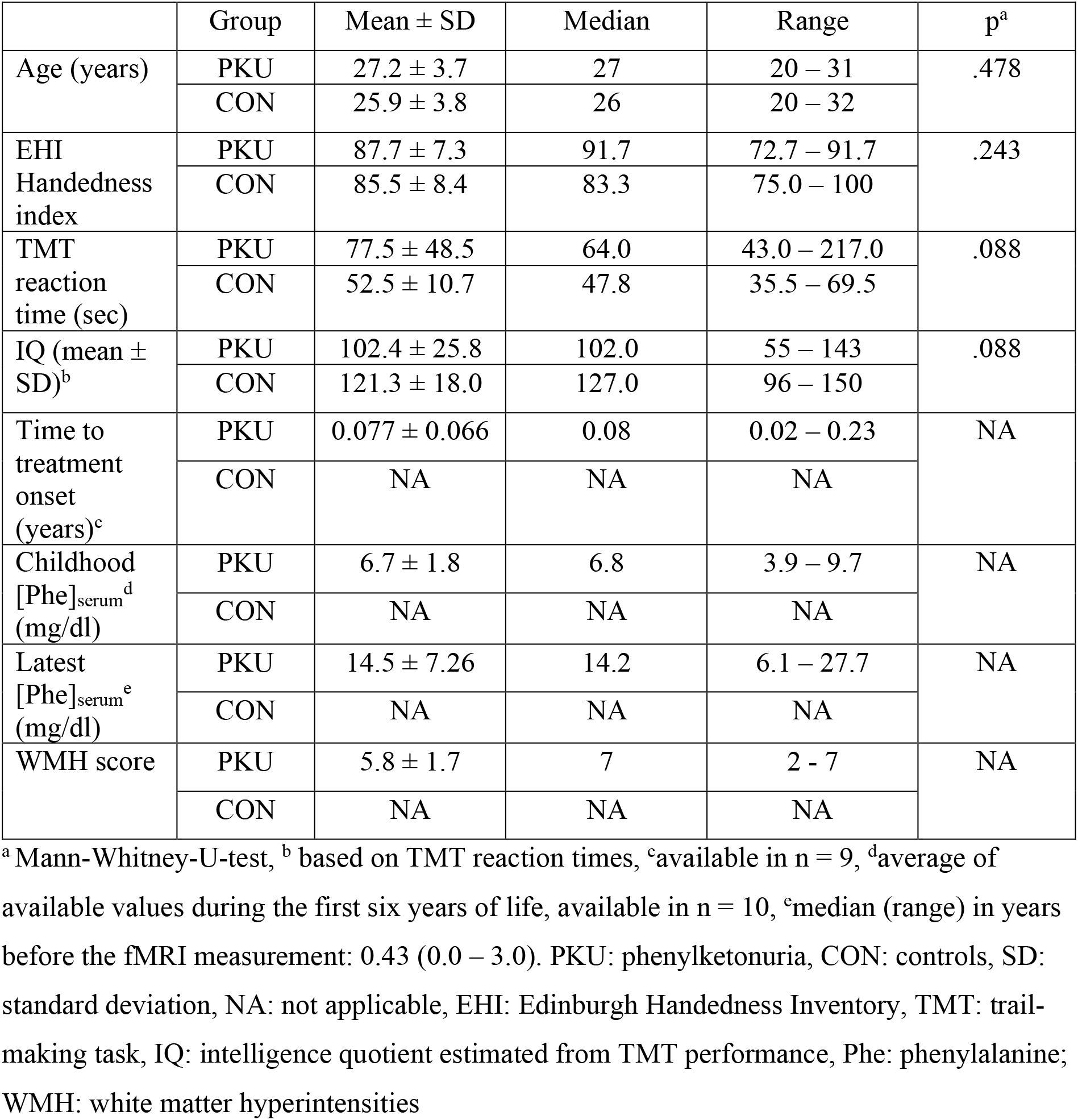
Demographical and clinical characteristics of female PKU patients (n = 11) and controls (n = 11)

| | Group | Mean ± SD | Median | Range | p<sup>a</sup> |
| --- | --- | --- | --- | --- | --- |
| Age (years) | PKU | 27.2 ± 3.7 | 27 | 20 – 31 | .478 |
| | CON | 25.9 ± 3.8 | 26 | 20 – 32 | |
| EHI Handedness index | PKU | 87.7 ± 7.3 | 91.7 | 72.7 – 91.7 | .243 |
| | CON | 85.5 ± 8.4 | 83.3 | 75.0 – 100 | |
| TMT reaction time (sec) | PKU | 77.5 ± 48.5 | 64.0 | 43.0 – 217.0 | .088 |
| | CON | 52.5 ± 10.7 | 47.8 | 35.5 – 69.5 | |
| IQ (mean ± SD)<sup>b</sup> | PKU | 102.4 ± 25.8 | 102.0 | 55 – 143 | .088 |
| | CON | 121.3 ± 18.0 | 127.0 | 96 – 150 | |
| Time to treatment onset (years)<sup>c</sup> | PKU | 0.077 ± 0.066 | 0.08 | 0.02 – 0.23 | NA |
|  | CON | NA | NA | NA |  |
| Childhood [Phe]<sub>serum</sub><sup>d</sup> (mg/dl) | PKU | 6.7 ± 1.8 | 6.8 | 3.9 – 9.7 | NA |
|  | CON | NA | NA | NA |  |
| Latest [Phe]<sub>serum</sub><sup>e</sup> (mg/dl) | PKU | 14.5 ± 7.26 | 14.2 | 6.1 – 27.7 | NA |
|  | CON | NA | NA | NA |  |
| WMH score | PKU | 5.8 ± 1.7 | 7 | 2 - 7 | NA |
|  | CON | NA | NA | NA |  |

<sup>a</sup> Mann-Whitney-U-test, <sup>b</sup> based on TMT reaction times, <sup>c</sup>available in n = 9, <sup>d</sup>average of available values during the first six years of life, available in n = 10, <sup>e</sup>median (range) in years before the fMRI measurement: 0.43 (0.0 – 3.0). PKU: phenylketonuria, CON: controls, SD: standard deviation, NA: not applicable, EHI: Edinburgh Handedness Inventory, TMT: trail-making task, IQ: intelligence quotient estimated from TMT performance, Phe: phenylalanine; WMH: white matter hyperintensities

### Acquisition and analysis of MRI data

MRI data were acquired at 3 Tesla including rs-fMRI (9:45 min of wakeful rest, eyes open), T1-weighted 3D data as reported previously [29], and 2D T2-weighted fluid-attenuated inversion recovery (FLAIR) [12]. WMH in PKU were visually scored on FLAIR images by a radiologist (BS) [36].

fMRI data preprocessing was identical with the FAS study [29] and was mainly based on fMRIPrep [37] (RRID:SCR_016216) with subsequent denoising [38]. MRI data quality control comprised visual inspection of structural data and preprocessing reports as well as exclusion of subjects with excessive head motion based on motion parameter estimates and visual quality indicators identical to the FAS study [29]. Head motion did not differ significantly between groups (see Table 2).

**Table 2.** Group comparison of head motion estimates. Data derived from preprocessing (final sample after exclusion of subjects with excessive head motion).

|  | PKU<sup>a</sup> | CON<sup>a</sup> | p<sup>b</sup> |
| --- | --- | --- | --- |
| Mean FD (mm) | 0.167 (0.10 – 0.28) | 0.150 (0.12 – 0.25) | 0.898 |
| Maximum FD (mm) | 0.865 (0.25 – 1.75) | 0.946 (0.55 – 1.65) | 0.300 |
| Percentage of motion spikes | 1.282 (0.00 – 8.97) | 1.709 (0.43 – 6.84) | 0.519 |

<sup>a</sup> median (range) <sup>b</sup>Mann-Whitney-U-test result. PKU: phenylketonuria, CON: controls, FD: framewise displacement

FC analyses were based on 400 atlas regions [39] matched to 17 subnetworks [40]. FC between regions in 10 cognition-related subnetworks [29] was calculated [41]. As a basis for subsequent modelling, we calculated multiple linear regression models (one model per pair of regions), comparing z-transformed correlation coefficients (dependent variable) among regions of interest between PKU subjects and controls. The following independent variables were included: group (PKU vs. control, categorical), age (normalized to center: 0, and standard deviation: 1), mean framewise-displacement (FD), and a constant term (intercept). Main effects of the factor group (PKU vs. controls, two-sided) were the basis of the subsequent analyses at different spatial scales, analogously to the FAS study [29]: The subsequent hierarchical analysis comprised the following steps: first, determining whether there is any alteration of FC in PKU subjects compared with controls in the full cognitive connectome globally (using HC) and subsequently aiming to resolve these findings: at the level of either FC within each network or between-network (concatenated regions per network) connectivity (using HC), and finally at the most detailed level of individual connections (conventional mass-univariate analysis with false-discovery rate (FDR) adjustment [42] for multiple comparisons). Please refer to the previous study for further model and software implementation details [29].

## Results

We observed altered FC of cognition-related brain regions in this sample of PKU patients compared with controls in the global analysis.

HC global testing revealed at least rare and weak group differences within this high dimensional dataset (Fig. 1B). The global null hypothesis for between-network connectivity could not be rejected (Fig. 1C).

**Fig. 1.**
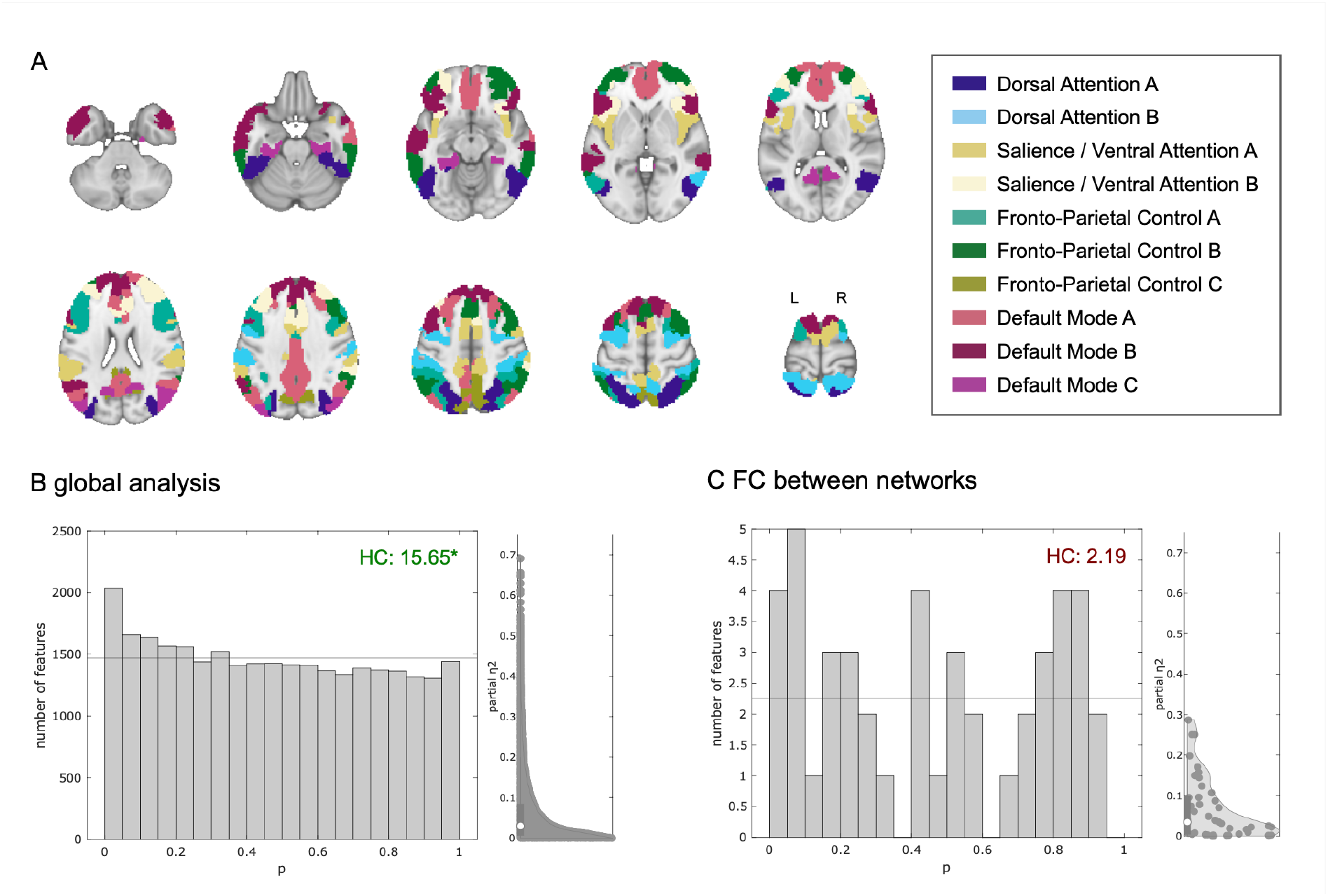
Global analysis and between-network functional connectivity differences of cognition-related brain networks. A) Overview of the ten cognition-related sub-networks based on the cortical atlas used in this analysis (previously published in [29] under a creative commons license: http://creativecommons.org/licenses/by/4.0/). B) P-value histogram and half violin plot of standardized effect size estimates of multiple individual linear models (main effect of group) comparing functional connectivity among all 243 single regions between PKU patients and control subjects. Under the null-hypothesis of equal functional connectivity in both groups, equal numbers of p-values are expected in each histogram bin (horizontal line). The histogram shows an excess of low p-values. The existence of at least rare and/or weak effects visualized in the histogram is confirmed by a quantitative test of the joint hypothesis based on higher criticism statistics. This means that regarding a significant number of functional connections, PKU patients differ from healthy controls. C) The null-hypothesis could not be rejected in the between-network connectivity analysis. HC: higher criticism test statistic (green: global null hypothesis rejected; red: global null hypothesis not rejected)

Group differences in FC were observed within 6 out of 10 cognition-related networks (Fig. 2). Based on the HC statistic, differences were most pronounced in 2 out of 3 subcomponents of the DMN (A and C; no effects observed in subnetwork B). Beyond, we found effects within all 4 attention-and salience-related subnetworks. In contrast, no effects were observed within the three subcomponents of the fronto-parietal control network.

**Fig. 2.**
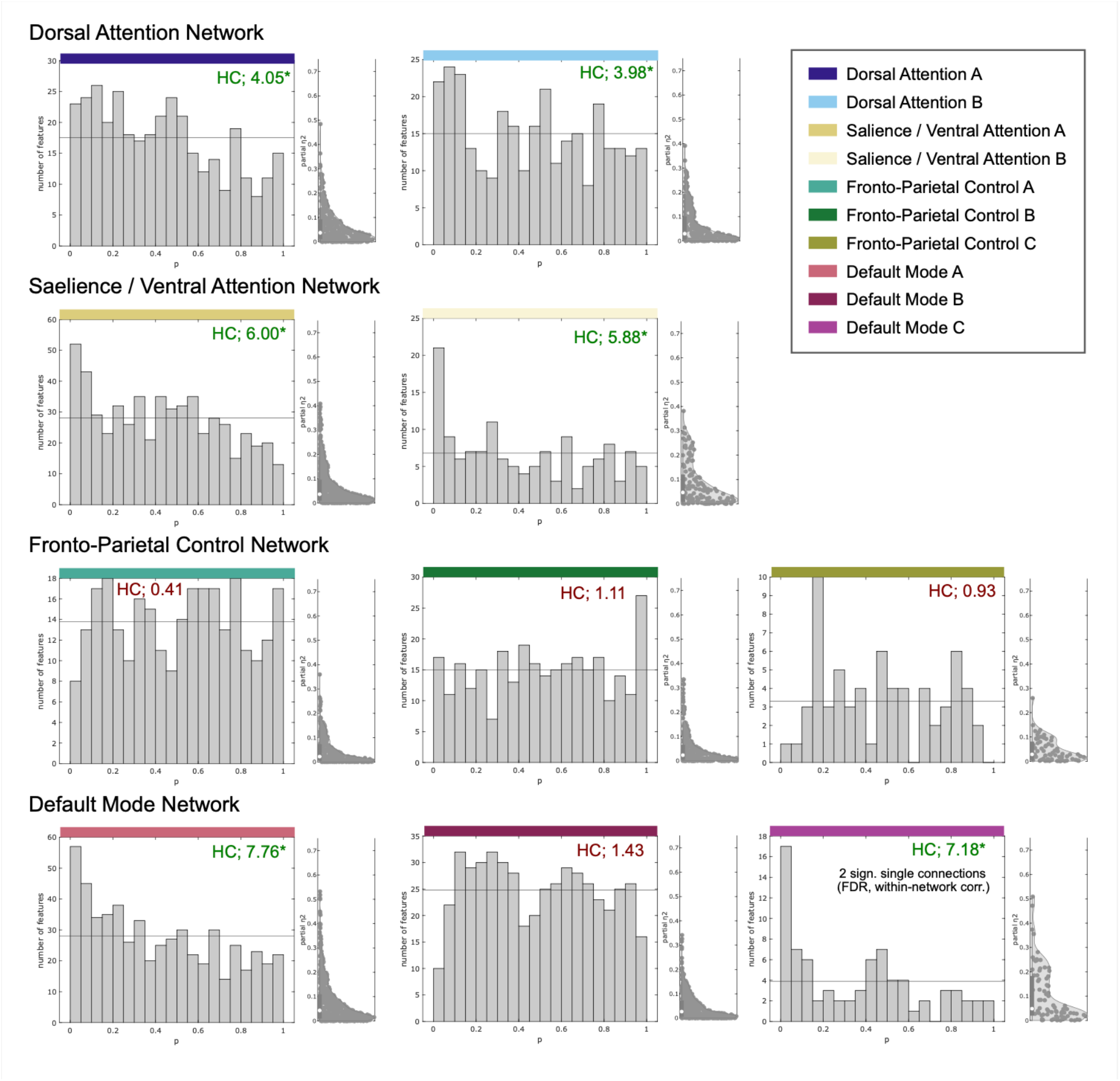
Within-network functional connectivity differences in cognition-related brain networks. Six (out of ten) sub-networks exhibited altered functional connectivity in PKU patients compared with controls (p-value histograms and half violin plots of standardized effect size estimates): All four attention-related sub-networks and two sub-networks of the default mode network exhibited an excess of low p-values of multiple linear models (main effect of group) compared with the expected number under the global null hypotheses (horizontal line). This means that the PKU patients differed from healthy controls regarding at least rare and/or weak effects within these networks. HC: higher criticism test statistic (*green: global null hypothesis rejected; red: global null hypothesis not rejected)

Two single connections within the DMN C sub-network exhibited significant group differences with FDR-adjustment within this network only. These findings did, however, not survive FDR-correction across all connections.

## Discussion

Using HC-based methodology, we were able to observe alterations of FC in young adult females with early-treated PKU when compared to healthy individuals. Despite the small sample size, results follow an interpretable pattern which show group differences attributable to attention-related functional networks and the DMN.

The involvement of the DMN is in line with limited previous fMRI results [12, 17]. The observed involvement of only the 2 DMN subnetworks (A and C) with substantial share of posterior brain regions corresponds to the typical distribution of WMH [9] as well as previous morphometric findings [10, 11, 43]. By visual comparison of atlas regions, the DMN A subnetwork substantially overlaps with the previously proposed DMN midline core, while the DMN C subnetwork mainly overlaps with the DMN medial temporal lobe subsystem [44].

To date, there is no previous fMRI evidence of altered attentional systems in PKU. However, this observation is in line with the neuropsychological profile (i.e. involving impairments in visual-spatial attention, and sustained attention,) reported in adults with early treated PKU [7, 8]. In this study, group differences appear pronounced in the salience / ventral attention networks compared with the dorsal attention network.

The negative finding in the fronto-parietal control network appears surprising in the context of the classical assumption of prefrontal specificity of deficits in children with PKU [45, 46]. Executive control deficits have also been reported in adults with PKU as part of a wider spectrum of neurocognitive deficits [4-6, 8]. The absence of findings in the fronto-parietal networks in this study might, however, correspond with a potential regression of certain PKU-related deficits during aging [7, 47-49]. It is also in line with previous task-based fMRI studies with negative [13] or only weak findings [12, 15] in fronto-parietal control networks in adults.

The comparatively straightforward interpretability of findings and correspondence with previous knowledge might indicate a potential benefit of a network-wise HC analysis in rare diseases, to which the previously discussed limitations (such as small sample sizes) apply.

In conclusion, this sample exhibited FC alterations in networks which are in line with limited previous neuroimaging findings and the clinical neuropsychological profile in adult PKU patients.

## Limitations

Application of HC to FC-analyses in this or similar ways [29, 50, 51] is a relatively novel approach with limited knowledge compared with conventional mass-univariate fMRI analyses. However, there is conceptual overlap with the more established method of network contingency analysis [52]. Methodological limitations have been discussed in detail in our related FAS article [29].

The generalizability of our observations is limited by the small sample size, restriction to women in a relatively narrow age range and IQ differences between PKU and controls, as well as by a limited clinical, neuropsychological, and social characterization of the sample. Beyond, sample size and statistical power restrict the ability to identify which exact connections were altered and how exactly they were affected. Our subnetwork-level results might, however, be informative for planning and interpreting future larger scale neuroimaging studies in PKU, e.g. by narrowing down hypotheses to the affected subnetworks observed here.

## Data Availability

Due to German data protection regulations and to safeguard subject confidentiality, data on the level of individual subjects cannot be made available

## List of abbreviations

DMN: default mode network
FAS: fetal alcohol syndrome
FC: functional connectivity
FD: frame-wise displacement
FLAIR: fluid-attenuated inversion recovery
fMRI: functional magnetic resonance imaging
HC: higher criticism
IQ: intelligence quotient
MRI: magnetic resonance imaging
OMIM: Online Mendelian Inheritance in Man
PKU: phenylketonuria
rs-fMRI: resting state functional magnetic resonance imaging
TMT: trail-making task
WMH: white matter hyperintensities

## Declarations

### Ethics approval and consent to participate

This study was approved by the ethics committee of the University of Münster and the Westphalian Chamber of Physicians in Münster (reference number: 2012-314-f-s with amendment). All study procedures were carried out after obtaining written informed consent and in accordance with the Declaration of Helsinki.

### Consent for publication

Participants provided informed consent for publication of study results. Consent for publication of individual data: not applicable (no individual data reported).

## Availability of data and materials

On request to the authors, further intermediate data on a level independent from the individual subjects can be shared. Due to German data protection regulations and to safeguard subject confidentiality, data on the level of individual subjects cannot be made available (no participant consent for sharing these primary data).

Software and statistical algorithms used for these analyses are available as referenced in the methods section and the more detailed methods description in our previous related work [29]. Further in-house code was used for individual aspects of data handling only and could be made available to other researchers on reasonable request.

## Authors’ contributions

BS, BP, RF, JW conceived and designed the study. BS, RF, JR, SG, BP collected the data. BS and BP carried out main data analyses. CM, SG, JR contributed to or supervised parts of the data analysis. BS, AM, BP, CM, RF, JR interpreted the results. AM, JR, SG, BS carried out main literature research. BS drafted the manuscript. All authors contributed to the manuscript for important intellectual content, approved the final manuscript and agree to take responsibility according to the journal guidelines.

## Acknowledgements

We thank all participants for taking part in this study. We acknowledge the kind support of staff at the Translational Research Imaging Center (University Hospital Muenster, Clinic of Radiology), Agnes van Teeffelen-Heithoff at the metabolic laboratory (University Hospital Muenster, Department of Pediatrics). We thank Mahboobeh Dehghan Nayyeri and Niklas Wulms for assistance with BIDS data handling [53]. The color scheme representing networks in Fig. 1 has been inspired by Paul Tol’s guide on accessible color schemes (https://personal.sron.nl/~pault/data/colourschemes.pdf). Half-violin plots were generated with Bastian Bechtold’s Matlab plugin (https://github.com/bastibe/Violinplot-Matlab, DOI: 10.5281/zenodo.4559847). ChatGPT (GPT3.5, http://chat.openai.com/) was used for minor programming assistance (suggestions for efficient Matlab code structure). After using this tool, the author (BS) reviewed the output and takes full responsibility for the resulting analyses. Negative results from a conventional FC analysis in the cognitive control network [54] in a preliminary subsample from this study have been presented at the 2014 annual meeting of the Organization for Human Brain Mapping (OHBM). Main results of this HC-based analysis were presented at the 2024 OHBM annual meeting [55].

